# Psychometric properties of Persian-version PTSD questionnaire Applied through Phone Follow-Ups: An experience in PERSIAN Traffic Cohort

**DOI:** 10.1101/2025.09.06.25334907

**Authors:** Nasrin Shahedifar, Alireza Razzaghi, Mohammad Asghari Jafarabadi, Mostafa Farahbakhsh, Shahrzad Bazargan-Hejazi, Mina Golestani, Leili Abedi Gheshlaghi, Homayoun Sadeghi-Bazargani

## Abstract

**Aims:** Traumatic events of road traffic accidents (RTAs) lead mostly to post-traumatic stress disorders (PTSD). We aimed to investigate the psychometric properties of the PTSD-5 checklist at consecutive phone follow-ups and its applicability in screening for PTSD in RTA survivors.

**Subject and Methods:** The study assessed the reliability and applicability of PTSD-5 tool through phone surveys, within a national traffic cohort platform. We recruited 150 patients at one-, six-, and twelve-month follow-ups after discharge, enrolling 50 patients per phase. We measured reliability and the agreement using Kappa coefficients, percentages of agreement and Bland-Altman method. Data were analysed using STATA statistical software package version 15.

**Results:** The participants were mostly male (79.8%), with a mean age (SD) of 41 (14.7). More than 70% of participants were married or employed or educated. It had excellent internal consistency reliability at total and follow-up levels (Cronbach’s α = 0.95) and good total stability reliability (ICC=0.77,95%CI:0.68, 0.83). One month follow-up data showed poor stability reliability (ICC=0.32,95%CI:-0.10, 0.59). The agreement between test and retest data at six- and 12-month follow-ups was excellent. Total Kappa coefficient and the percentage of agreement were 0.58 and 71.3%, respectively. The Bland Altman plot on one-year follow-up data revealed more agreement at healthier state (lower scores).

**Conclusion:** The Persian PTSD checklist was highly reliable and applicable through phone interviews post discharge in road traffic injury adult patients. It was suitable to apply within the national cohort study and would decline the data collection costs.

## Introduction

Each year, almost 50 million people experience traumatic event through road traffic accidents (RTAs) [1]. Post injury, of its physical, psychological and social outcomes, post-traumatic stress disorder (PTSD) often follows a traumatic event. PTSD triggers major distress, functional impairment, poor quality of life, and interferes with role function (performance at school/ work/ homemaking, relationships with family/ friends) [2, 3]. As defined by American Association of Psychology, PTSD, a kind of anxiety disorder experienced subsequent to a stressful threatening or disastrous condition, is accompanied by recurrent remembering of an event in thoughts and dreams, avoiding situations which induce trauma memories and severe arousal [4].

In low and middle income countries (LMICs), particularly in Iran, road traffic crashes (RTCs) cause the second most deaths [5, 6] and impair patients’ quality of life and daily functioning [7, 8]. Witnessing accident scenes, death and severe stress that are faced in the blink of an eye, would severely impact a witness’s psychological dimension and create a permanent fear and apprehension or depression [9]. The prevalence of probable PTSD among road traffic injuries (RTIs) survivors was high (15.4%-39.2%) [2, 3, 9-11]. However, survivors’ mental health and status are less concerned than their physical aspect [12, 13], particularly after discharge. To achieve as valid results as expensive psychiatry evaluation, we need to apply a reliable tool in screening patients’ PTSD post discharge. Moreover, it should be remotely completed via phone interviews, regarding patients’ physically limited situation after injury as well as rare opportunity to have in-person visits, in some settings especially in LMICs. And the screening must be done in consecutive follow-up times through a year post injury. Most often, an appropriate circumstance to run them all in a single research is rarely provided for researchers. However, we could manage to conduct such research through a national cohort platform of Post-crash Traffic Safety and Health Cohort Study, <u>P</u>rospective <u>E</u>pidemiological <u>R</u>esearch <u>S</u>tudy in Ir<u>AN</u> (PERSIAN) [14]. The cohort study provided us with a great opportunity to apply the tool and assess its psychometric properties via phone interviews and through three times post discharge. Then, to prevent psychosocial problems post-injury from becoming chronic, subsequent to approval of its reliability, we would apply the tool to screen all RTI hospitalized patients during their first year after injury at various follow-up phases.

One of the most common tool to screen patients’ PTSD is primary care PTSD screen checklist (PTSD-5) for the Diagnostic and Statistical Manual of Mental Disorders, Fifth Edition (DSM-5). That could be administered with great care and sensitivity. As we have to use a complete globally approved tool and short enough to fill out in phone surveys and short time, we utilized and psychometrically assessed PTSD-5 through phone interviews at three follow-up phases, namely one, six, and twelve months after injury (1, 2, and 3 F-up, respectively). Also, to our knowledge, the paucity of psychometric study of such all-inclusive tool on creating the health profile of RTCs survivors is sensible reason to examine internal consistency reliability and stability reliability of PTSD among clinical population through phone surveys at three phases post injury.

## Materials and Methods

### Assessment of reliability

The tool’s psychometric properties was examined at three times after crash through phone interviews among a clinical population. The RTIs survivors recruited from the post-crash cohort study were the study source population [14]. All participants hospitalized in one of two referral trauma centers, were followed up in the cohort study at 1, 2, and 3 F-ups post injury. Since 2019, all hospitalized patients have been recorded in the database of Integrated Road Traffic Injury Registry System (IRTIRS) [15]. We took 50 patients at each follow-up, because to evaluate the reliability, the least sample number was 50 for detecting the value of 0.4 for ICC, with alpha at 0.05 and power defined at lower than 90% [16]. Then, patients were randomly recruited from those who must be called via phone to complete the questionnaire at 1, 2, and 3 F-ups. Each phases’ participants were different from others, then we had 150 patients in total who took part in test and retest phases between 21 May 2020 and 15 June 2020. The test and retest were conducted with a time span of averagely 10 days (from 9 to 11 days). The subjects did not have any treatment or changes in their medication during the time between test and retest. As inclusion criteria, participants must be hospitalized due to RTIs for at least 24hrs, registered, aged 18 or over, and informed consent was requested. Exclusion criteria for this step included having any disorders like psychiatric problems that may affect the validity of responses or a disastrous event experienced during the test-retest interval (e.g. disease, another car crash, fall and so on). A trained interviewer with more than four-year experience routinely collected data from registered patients on phone calls [17]. As the interviewer filled them out, the completeness of tools was mostly guaranteed. The selected participants were replaced by the same sex and age if they refused to participate.

### Measures

#### PTSD questionnaire

Posttraumatic Stress Disorder (PTSD) as a common, chronic and weakening disorder, should be examined after an injurious event. The brief tool of Primary Care-PTSD-5 has been broadly applied to screen for PTSD since it has excellent diagnostic accuracy and clinical utility]18[. The PC-PTSD-5 includes five dichotomous items requiring “yes” or “no” responses to assess post-traumatic symptoms over the past month and the total score ranged from 0 for “no problem” to 5 for “with PTSD symptoms”. A total score is calculated by the number of “Yes” in the five items [19]. The threshold for clinically significant PTSD symptoms was recognized by score three and more. The PC-PTSD score was calculated for test and retest data of all 150 participants, and separately at each follow-up phase (n=50).

#### Socio-demographic data

Basic socio-demographic data and selected crash-related information including used vehicle, counterpart vehicle, crash mechanism, and injured person’s role were collected.

### Statistical analysis

Initial analyses of the baseline characteristics and the tool data were accomplished using descriptive statistics. To compare the psychometric properties of the tool, we assessed internal consistency reliability and stability reliability. To assess the agreement of test and retest scores, we estimated Kappa agreement coefficients and Bland-Altman plot. Data were analysed using STATA statistical software package version 15 (StataCorp LLC, Texas) and a significance level of 0.05 was adopted.

#### Scale score reliability

### Internal consistency reliability

The extent to which the items in a scale are correlated is specified by internal consistency. The Cronbach’s Alpha statistic evaluates the homogeneity [20]. A value from 0.5 to 0.75 indicates moderate reliability, values between 0.75 and 0.9, and greater than 0.90 specify good and excellent reliability, respectively [21].

### Test–retest reliability

The repeatability assessment was measured using the test–retest method. Comparing the results of both responses looked at the consistency over time. To examine the correlation, we applied tau Kendal b and Spearman-Brown correlation coefficients. Acceptable Spearman-Brown coefficient is greater than 0.7 [22]. The Kappa agreement is a robust method in reliability testing. Its values represent no agreement (≤ 0), none to slight (0.01–0.20), fair (0.21–0.40), moderate (0.41– 0.60), substantial (0.61–0.80), and almost perfect agreement (0.81–1.00) [23]. The reliability of the method was assessed by intra-class correlation coefficient (ICC). In estimation of the intra-class correlation coefficient with 95% confidence intervals, we adopted the “single-rater type (k=1), absolute-agreement, 2-way mixed-effects model [21]. The measurement stability is evident by values >=0.75. Fair to good reliability is shown by ICC from 0.4 to 0.75, and the ICC of <0.4 points to poor reliability [24].

### Agreement

Besides, regarding restrictions of ICC and correlation coefficient in assessing the agreement between the tool’s test and retest data [25], Bland-Altman method was applied to examine their agreement. The result is a scatter plot XY, in which the Y axis displays the difference between the two paired measurements (A-B) and the X axis indicates the average of these values ((A+B)/2) [26]. Thus, its vertical and horizontal axes indicate difference between, and average of test and retest scores, respectively (Figure 2). The limits of agreement were also estimated as mean difference ±1.96 standard deviations of the difference. A good agreement between test and retest scores would be represented by a near to zero mean difference and with a maximum of 5 % or less points residing outside the limits of agreement [27].

**Figure 2.**
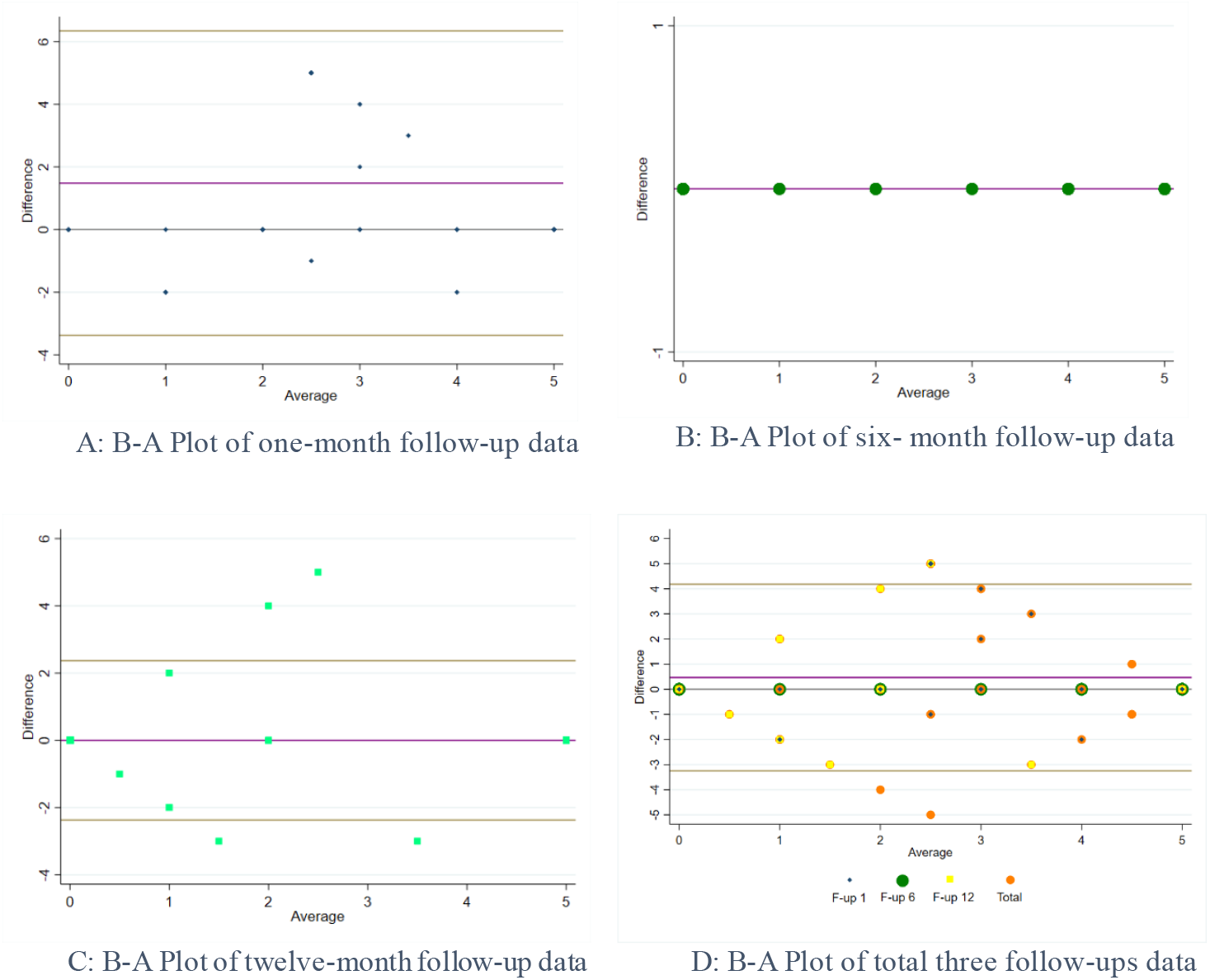
Bland-Altman plots: agreement between total test and retest scores and test and retest scores at three follow-ups. Y axis, difference between test and retest scores; X axis, average of test and retest scores

### Floor and ceiling effects

The floor and ceiling effects were acquired from the percentage of the lowest and the highest scores responded for individual items of the tool. A ceiling effect is present when more than 15 % of responses attributed to the highest score, [27]. Ceiling and floor effects are noticed as moderate and substantial when up to 25% and higher than 25%, respectively [28].

## Results

### Sample characteristics

The study was conducted among 120 (79.87%) male and 30 (20) female participants. The total mean age (SD) was 41 (14.7) years old. The majority of participants were adults between 25-64 years old, and more than 70% of respondents were married, educated, and employed. The vehicle-vehicle collision (50%-56%) was the main crash mechanism followed by Vehicle-pedestrian collision. The most participants at each follow-up phase were motorcycle and car drivers (64%-52%) and Passenger/ pillion passenger (24%-32%).

### Scale score reliability

#### Floor and ceiling effects

The proportion of patients with the lowest (0) and highest (5) scores were 32% and 48% (n=150). There were reported floor and ceiling effects at each follow-up (F-up1: 16, 60; F-up2: 36, 38; F-up3: 44, 46).

#### Internal consistency reliability

Cronbach’s alpha coefficient

For the total test data (N=150), the overall Cronbach’s α coefficient resulted in 0.95 showing excellent internal consistency. The Cronbach’s α coefficients were assessed for each follow-up phase (N= 50) as 0.92, 0.94, and 0.98 at one, six, and 12 month after crash, respectively.

#### Stability reliability

Testing the test-retest data correlation, the Spearman-brown correlation coefficients were 0.39, 1.00, and 0.93 at one, six, and 12 months after crash, respectively, as well as 0.78 for total data.

The test-retest reliability of the tool was examined by the ICC. One month follow-up data showed poor stability reliability. The six- and 12-month follow-ups’ ICC showed the excellent absolute agreement between test and retest data. For total data, the coefficients suggested good reliability and stability (Fig. 1). The minimum total ICC at item levels was 0.73 for question two.

#### Test–retest reliability

The total Kendall’s tau-b of 0.59 presented the correlation between total test and retest data (N=150). At follow-up level, it ranged from 0.15 for F-up 1 to 1.00 for F-up 2, with P<0.0001 (Table 2). The kappa agreement coefficients specified fair to perfect ranged from 0.38 at F-up 1.00 to 1.00 at F-up 2, with P<0.0001. The percentage of agreement was high enough [27] (Table 2).

**Table 1.**
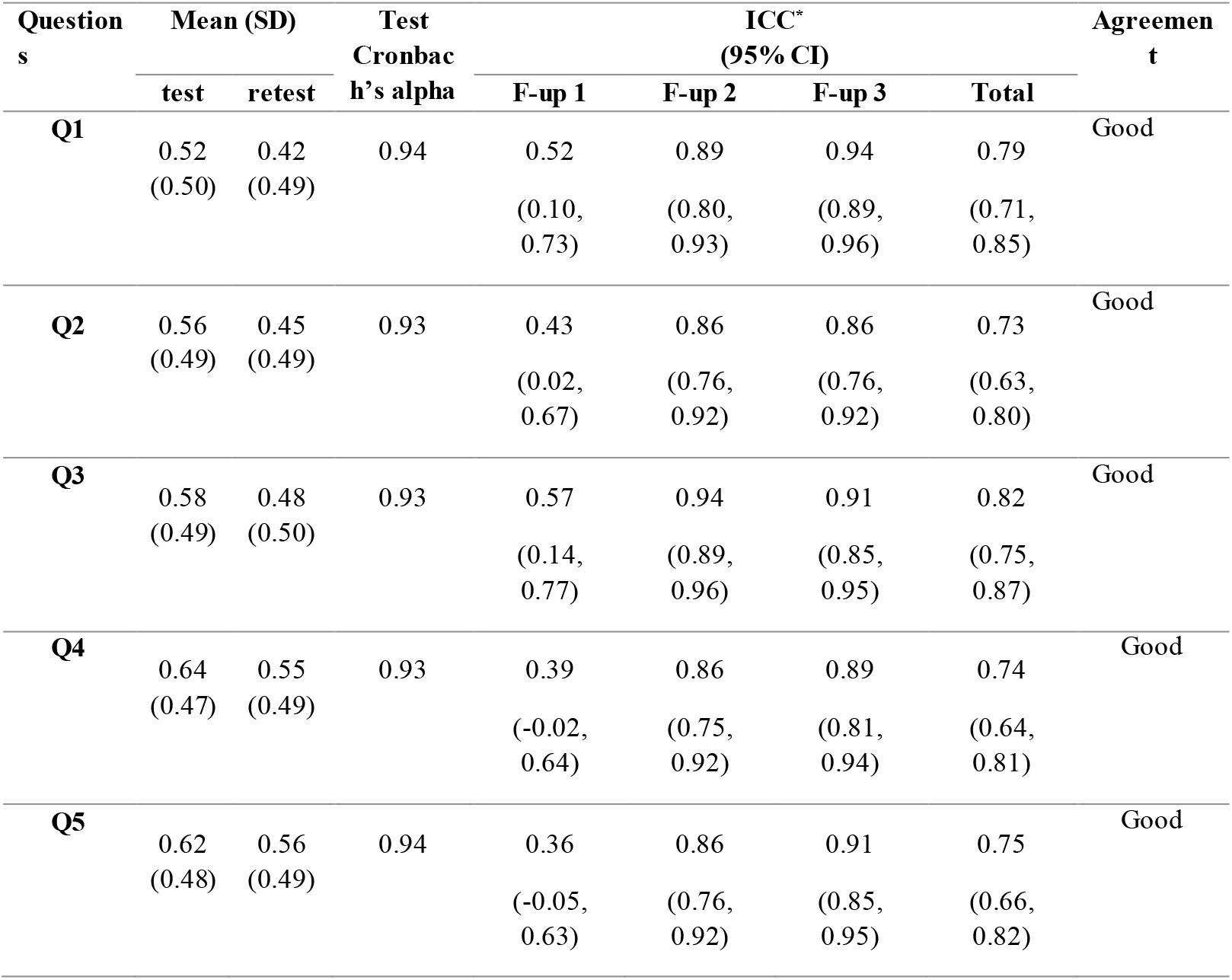
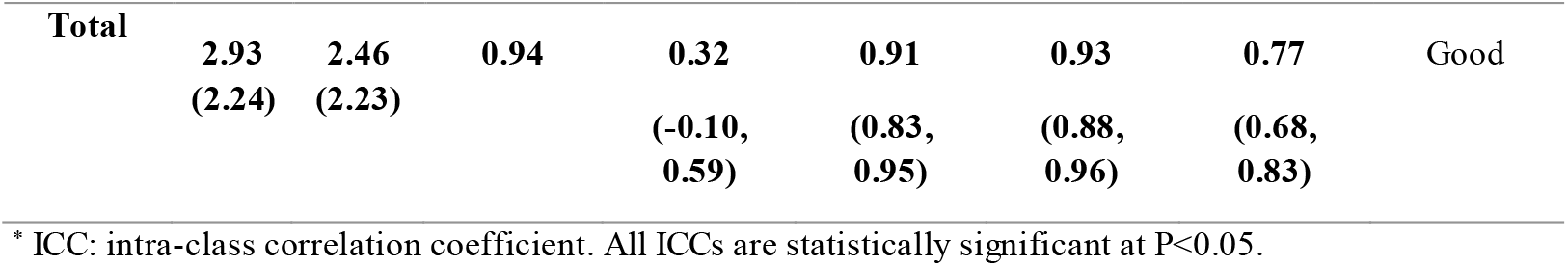
Reliability indices of the Persian version of PTSD questionnaire in phone survey (N=150)

**Table 2.**
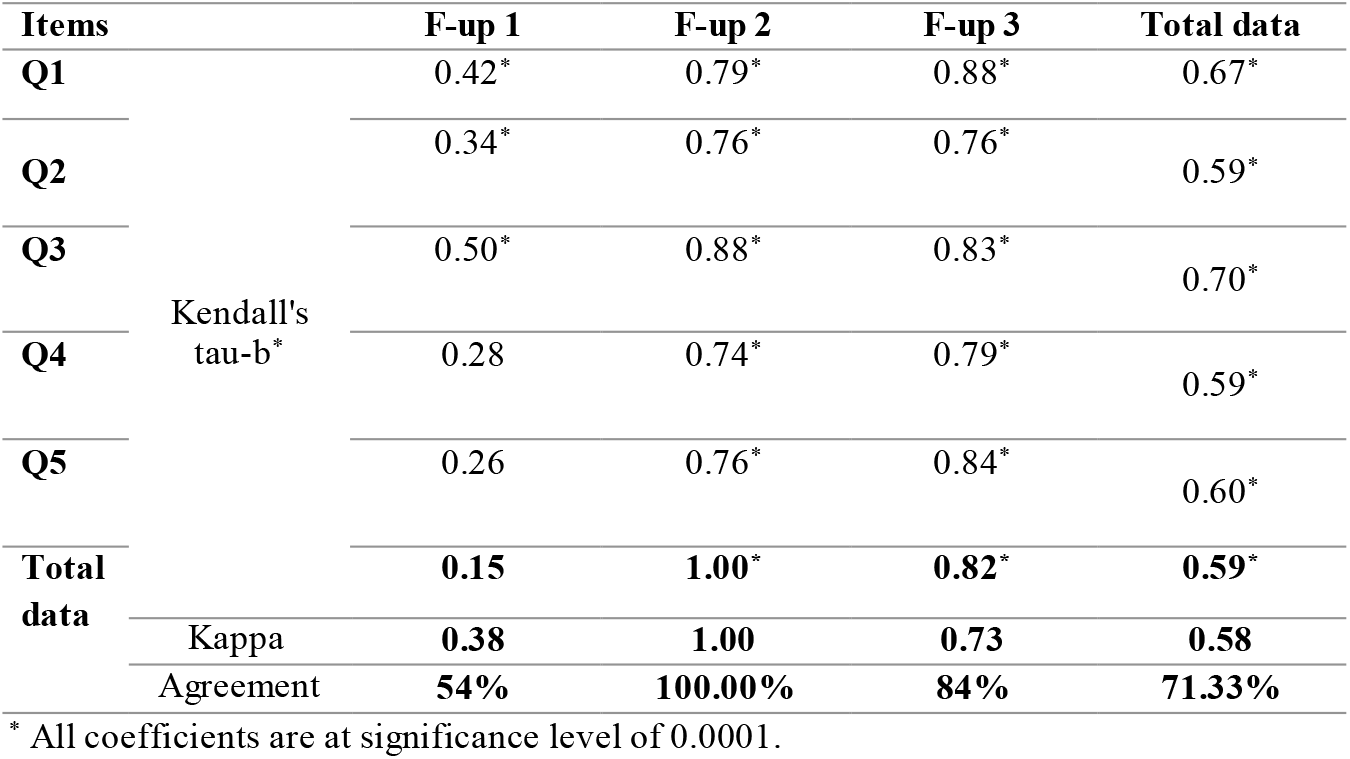
Test-retest correlation and agreement of the Persian PTSD at three phone follow-ups.

Bland-Altman plot

At first follow-up, 95% of points lie in limits of agreement (−3.38, 6.34). The great density and agreement is around the cut point of three. The B-A plot, at six month follow-up, shows full agreement between test and retest scores. After one year, the test and retest scores of the questionnaire indicated more agreement at lower scores, i.e. healthier state. The agreement between total scores of test and retest data collected from 150 participants has been reached as the majority of points fall around the mean difference (0.47) and 95% of points are shown at the limits of agreement (−3.24, 4.18).

## Discussion

The study examined the psychometric properties of the available Persian version of PTSD-5 via phone interviews at three different times post event as the special features of the work. Tools’ psychometric properties varies based on its data-collection technique and a population source [29]. Regarding the lack of such assessment in phone interviews and among road traffic injuries which are the second cause of death in Iran [5, 6], we prepared this work. The opportunity of running such study on a post-crash cohort platform was a major reason, too. The study revealed that Persian PTSD-5 has excellent internal consistency reliability and great stability reliability in phone-based assessment.at six and twelve months post injury. The study showed evidence of excellent internal consistency reliability compared with a study in China [30, 31].

The story of test and retest reliability at one month follow-up was different and perplexing, as it specified excellent internal consistency reliability, but poor stability reliability. An important item may affect the pattern of poor stability reliability would be pertinent to acute stress syndrome (ASD). Its four symptom clusters were re-experiencing, avoidance, increased arousal, and dissociative symptoms, which are alike to those of PTSD, except the last one [32]. The onset of ASD symptoms are within 4 weeks of the traumatic experience [32]. ASD is claimed to become the PTSD’s potential clinical predecessor [33]. Providing the continuation of symptoms for more than 4 weeks, the diagnosis is changed to PTSD [32-34]. In the current study, the test phase was conducted averagely 30 days after event, which mostly indicated symptoms of ASD. Then, the retest phase was done averagely 40 days post event in which symptoms of PTSD were reported, as ASD symptoms were lessened in patients without PTSD symptoms. The “Yes” response to three questions about avoidance, feeling detached, and feeling guilty were fewer in the retest than the test measurements.Accordingly, the findings are in complete consistency with the theory stated in DSM-V [34]. In fact, we could recognize trauma survivors, with high risk for PTSD, in the acute phase [35] at first-month follow-up.

For further investigation about poor stability reliability at one-month phone-based follow-up data, we specified that the screening tool revealed 36 and 19 patients with PTSD at test and retest phases, respectively, out of 50 patients followed up one month post injury. Following the phone-based screening interviews, next stage in the extensive PTC is defined to invite patients with PTSD to a psychiatry visit. Accordingly, 12 out of 36 patients included in this study were visited by our psychiatrist two to three months post injury and only two of them were recognized with PTSD (16.6%), four (33%) with major depressive disorder, and six (50%) with anxiety disorder without specific subtype.

The study examined the tool’s phone-based reliability at three different time spans (one, six, and twelve months) post event, as justified elsewhere [17]. Time since collision is a determining factor for PTSD [36]. The PTSD prevalence was 30.1% and 14.0% one month and one year post trauma among survivors of unintentional traumas [37], and 18.4% of RTI patients fulfilled the criteria for PTSD within 6 months [38]. Some studies revealed the occurrence of delayed-onset PTSD starting 6 months after a traumatic event among accident victims [39-41]. In order to shorten the time to remission of PTSD symptoms and hospitalization, timely recognition and intervention are essential [37, 38]. However, the treatment options are rarely focused on psychological outcomes post trauma [13]. Moreover, three time periods (one, six, and 12 months after injury) were included to investigate various patterns such as delayed-onset PTSD, on the basis of reasons obtained from expert panel experiences and other studies approved the necessity of screening and treating post-injury psychological comorbidities in a timely manner [42, 43].

We preferred a reliable, valid, short and simple tool to use within the national cohort study, along with other various measurements. So, the primary care PTSD-5 was an appropriate choice to apply in screening patients’ PTSD. Moreover, as studies revealed the elderly, uneducated or low-educated patients might have more troubles in discriminating the levels of multiple choice questions and remembering them with their corresponding number [44, 45]. The RTI victims are mostly lower educated and eight percent of victims are the elderly [46]. Hence, the short version PTSD screening checklist is preferable. Also, phone interviews as a short data gathering method become essential in long-term projects, since repeated connections with patients will be needed. In phone surveys, patients and interviewers do not have to travel to other place [47].

## Limitations

Because the severity of symptoms among patients experienced traumatic events becomes higher than those experienced non-traumatic events [9], extrapolation of findings to other types of injuries and patients, and children and adolescents must be made with more caution. The pattern of disorder in patients with disability may become different, too. Regarding similar and simultaneous symptoms of PTSD and ASD, it is recommended to conduct the reliability test at 40 days post injury through phone interview after declining symptoms of ASD. Moreover, extra research on patients and age groups, as no previous study considered its psychometric properties at various phone follow-ups after RTIs. Further evaluation of the measure is necessary in senior people via phone survey due to their possible hearing problems.

## Conclusion

### Conclusion

The study revealed that Persian PTSD was highly reliable to be applied through phone interviews at six and twelve months post discharge in road traffic injury patients at 18 or above. Based on findings, we would suggest to apply it at least forty days post injury. We realized that the tool is highly reliable and suitable to apply within the national cohort study. It helps decline the data collection costs since the data could be gathered remotely.

## Data Availability

the associated data could be available by a reasonable request from scientists by sending email to the corresponding author Dr. Homayoun Sadeghi-Bazargani. The study website is https://cohortsafety.tbzmed.ac.ir/.

## Ethical approval

The study respected ethical precepts in research and was performed consistent with the principles of the Declaration of Helsinki. It was evaluated and approved by the Ethics Committee of the Road Traffic Injury Research Center, Tabriz University of Medical Sciences (cohort ethics code: IR.TBZMED.REC.1398.543). An informed consent was required from all participants.

## Consent for publication

All authors read and approved the final manuscript to be published.

## Authors’ contributions

HSB conceived and designed the study, HSB and NS conducted the analyses and, ….interpreted the results drafted the manuscript contributed in the critical drafting and revising of the manuscript for important intellectual content. All authors read and approved the final manuscript.

## Acknowledgements

The authors would like to thank the participants in the study.

## Availability of supporting data

Data are available upon reasonable request. The interested researchers may contact Dr. Homayoun Sadeghi-Bazargani (PI). The applications will be reviewed upon approval by the research council and the regional ethics committee of Tabriz University of Medical Sciences. The study website is https://cohortsafety.tbzmed.ac.ir/.

## References

1. santé, O.m.d.l., et al., World Report on Road Traffic Injury Prevention. 2004: World Health Organization.

2. Cowlishaw, S., et al., Posttraumatic Stress Disorder in Primary Care: A Study of General Practices in England. Journal of clinical psychology in medical settings, 2020: p. 1–9.

3. Blanchard, E.B., et al., Psychiatric Morbidity Associated with Motor Vehicle Accidents. The Journal of Nervous and Mental Disease, 1995. 183(8): p. 495–504.

4. Azad Marzabadi, E. and S.M. Hashemi Zadeh, The Effectiveness of Mindfulness Training in Improving the Quality of Life of the War Victims with Post Traumatic stress disorder (PTSD). Iranian journal of psychiatry, 2014. 9(4): p. 228–236.

5. Azami-Aghdash, S., et al., Barriers to and Facilitators of Road Traffic Injuries Prevention in Iran; A Qualitative Study. Bull Emerg Trauma, 2019. 7(4): p. 390–398.

6. Bazargan-Hejazi, S., et al., The Burden of Road Traffic Injuries in Iran and 15 Surrounding Countries: 1990-2016. Arch Iran Med, 2018. 21(12): p. 556–565.

7. Gopinath, B., et al., Predictors of health-related quality of life after non-catastrophic injury sustained in a road traffic crash. Annals of Physical and Rehabilitation Medicine, 2020. 63(4): p. 280–287.

8. Yohannes, K., et al., Prevalence and correlates of post-traumatic stress disorder among survivors of road traffic accidents in Ethiopia. Int J Ment Health Syst, 2018. 12: p. 50.

9. Salehi-Pourmehr Hanieh, et al., Delayed PTSD Prevalence in Road Traffic Crashes: a systematic review and meta-analysis. Unpublished work, 2022.

10. Iteke, O., et al., Road traffic accidents and posttraumatic stress disorder in an orthopedic setting in South-Eastern Nigeria: a controlled study. Scandinavian journal of trauma, resuscitation and emergency medicine, 2011. 19: p. 39–39.

11. Shabani Zahra, et al., A systematic review and meta-analysis of the prevalence of posttraumatic stress disorder (PTSD) in road traffic accidents survivors. Unpubished work, 2022.

12. Khan, S. and S. Haque, Trauma, mental health, and everyday functioning among Rohingya refugee people living in short- and long-term resettlements. Social Psychiatry and Psychiatric Epidemiology, 2020.

13. Fekadu, W., et al., Incidence of Post-Traumatic Stress Disorder After Road Traffic Accident. Front Psychiatry, 2019. 10: p. 519.

14. Sadeghi-Bazargani, H., et al., PERSIAN Traffic Safety and Health Cohort: A Study Protocol on Post-Crash Mental and Physical Health Consequences. Journal of Injury Prevention, 2022.

15. Sadeghi-Bazargani, H., et al., Developing a National Integrated Road Traffic Injury Registry System: A Conceptual Model for a Multidisciplinary Setting. J Multidiscip Healthc, 2020. 13: p. 983–996.

16. Bujang, M.A. and N. Baharum. A simplified guide to determination of sample size requirements for estimating the value of intraclass correlation coefficient: a review. 2017.

17. Marin, S., et al., The protocol for validating phone interview tools on post-discharge consequences of road traffic injuries. Journal of Injury and Violence Research, 2020. 12(3).

18. Jung, Y.-E., et al., A Brief Screening Tool for PTSD: Validation of the Korean Version of the Primary Care PTSD Screen for DSM-5 (K-PC-PTSD-5). Journal of Korean medical science, 2018. 33(52): p. e338–e338.

19. Fung, H.W., et al., Using the Post-traumatic Stress Disorder (PTSD) Checklist for DSM-5 to Screen for PTSD in the Chinese Context: A Pilot Study in a Psychiatric Sample. Journal of Evidence-Based Social Work, 2019. 16(6): p. 643–651.

20. Andresen, E.M., Criteria for assessing the tools of disability outcomes research. Archives of physical medicine and rehabilitation, 2000. 81: p. S15–S20.

21. Koo, T.K. and M.Y. Li, A Guideline of Selecting and Reporting Intraclass Correlation Coefficients for Reliability Research. J Chiropr Med, 2016. 15(2): p. 155–63.

22. Abedzadeh–kalahroudi, M., et al., Psychometric properties of the world health organization disability assessment schedule II-12 Item (WHODAS II) in trauma patients. Injury, 2016. 47(5): p. 1104–1108.

23. McHugh, M.L., Interrater reliability: the kappa statistic. Biochem Med (Zagreb), 2012. 22(3): p. 276–82.

24. Cicchetti, D.V., Guidelines, criteria, and rules of thumb for evaluating normed and standardized assessment instruments in psychology. Psychological assessment, 1994. 6(4): p. 284.

25. Sadeghi-Bazargani, H., et al., Psychometric properties of the short and ultra-short versions of socioeconomic status assessment tool for health studies in Iran (SES-Iran). Journal of Clinical Research, 2016. 4.

26. Giavarina, D., Understanding Bland Altman analysis. Biochemia medica, 2015. 25(2): p. 141–151.

27. Yfantopoulos, J.N. and A.E. Chantzaras, Validation and comparison of the psychometric properties of the EQ-5D-3L and EQ-5D-5L instruments in Greece. The European Journal of Health Economics, 2017. 18(4): p. 519–531.

28. Anjos, D.B.M.D., et al., Reliability and construct validity of the Instrument to Measure the Impact of Valve Heart Disease on the Patient’s Daily Life. Revista latino-americana de enfermagem, 2016. 24: p. e2730–e2730.

29. Shahedifar, N., et al., Psychometric properties of the 12-item WHODAS applied through phone survey: an experience in PERSIAN Traffic Cohort. Health Qual Life Outcomes, 2022. 20(1): p. 106.

30. Cheng, P., et al., Psychometric Properties of the Primary Care PTSD Screen for DSM-5: Findings From Family Members of Chinese Healthcare Workers During the Outbreak of COVID-19. Frontiers in Psychiatry, 2021. 12.

31. Prins, A. and P. Ouimette, “ The primary care PTSD screen (PC-PTSD): Development and operating characteristics”.(vol 16, pg 257, 2003). Primary Care Psychiatry, 2004. 9(4): p. 151–151.

32. Randall D. Marshall, M.D. ; Robert Spitzer, M.D., and, and Michael R. Liebowitz, M.D., Review and Critique of the New DSM-IV Diagnosis of Acute Stress Disorder. American Journal of Psychiatry, 1999. 156(11): p. 1677–1685.

33. Nash, W.P. and P.J. Watson, Review of VA/DOD Clinical Practice Guideline on management of acute stress and interventions to prevent posttraumatic stress disorder. J Rehabil Res Dev, 2012. 49(5): p. 637–48.

34. Stein, D.J., et al., DSM-5 and ICD-11 definitions of posttraumatic stress disorder: investigating “narrow” and “broad” approaches. Depress Anxiety, 2014. 31(6): p. 494–505.

35. Bryant, R.A., et al., A review of acute stress disorder in DSM-5. Depress Anxiety, 2011. 28(9): p. 802–17.

36. Bedaso, A., et al., Prevalence and determinants of post-traumatic stress disorder among road traffic accident survivors: a prospective survey at selected hospitals in southern Ethiopia. BMC Emergency Medicine, 2020. 20(1): p. 52.

37. Santiago, P.N., et al., A systematic review of PTSD prevalence and trajectories in DSM-5 defined trauma exposed populations: intentional and non-intentional traumatic events. PLoS One, 2013. 8(4): p. e59236.

38. Frommberger, U.H., et al., Prediction of posttraumatic stress disorder by immediate reactions to trauma: a prospective study in road traffic accident victims. Eur Arch Psychiatry Clin Neurosci, 1998. 248(6): p. 316–21.

39. Utzon-Frank, N., et al., Occurrence of delayed-onset post-traumatic stress disorder: a systematic review and meta-analysis of prospective studies. Scand J Work Environ Health, 2014. 40(3): p. 215–29.

40. Chossegros, L., et al., Predictive factors of chronic post-traumatic stress disorder 6 months after a road traffic accident. Accid Anal Prev, 2011. 43(1): p. 471–7.

41. Guest, R., et al., Prevalence and psychometric screening for the detection of major depressive disorder and post-traumatic stress disorder in adults injured in a motor vehicle crash who are engaged in compensation. BMC Psychol, 2018. 6(1): p. 4.

42. Papadakaki, M., et al., Psychological distress and physical disability in patients sustaining severe injuries in road traffic crashes: Results from a one-year cohort study from three European countries. Injury, 2017. 48(2): p. 297–306.

43. Smid, G.E., et al., Delayed posttraumatic stress disorder: systematic review, meta-analysis, and meta-regression analysis of prospective studies. J Clin Psychiatry, 2009. 70(11): p. 1572–82.

44. Liang, Z., et al., Health-related quality of life among rural men and women with hypertension: assessment by the EQ-5D-5L in Jiangsu, China. Quality of Life Research, 2019. 28(8): p. 2069–2080.

45. Yang, F., et al., Do Rural Residents in China Understand EQ-5D-5L as Intended? Evidence From a Qualitative Study. PharmacoEconomics - Open, 2021. 5(1).

46. Sadeghi-Bazargani, H., et al., Epidemiology of Road Traffic Injury Fatalities among Car Users; A Study Based on Forensic Medicine Data in East Azerbaijan of Iran. Bull Emerg Trauma, 2018. 6(2): p. 146–154.

47. Shahedifar, N., et al., Psychometric properties of EQ-5D-3L applied at three phone-based follow-ups: an experience in PERSIAN cohort study. Bulletin of Emergency And Trauma, 2022: p. Submitted.

